# A genetic risk score improves risk stratification for anticoagulation-related intracerebral hemorrhage

**DOI:** 10.1101/2022.10.24.22281463

**Authors:** Ernst Mayerhofer, Livia Parodi, Savvina Prapiadou, Rainer Malik, Jonathan Rosand, Marios K Georgakis, Christopher D Anderson

## Abstract

**Background:** Intracerebral hemorrhage (ICH) is the most devastating adverse outcome for patients on anticoagulants. Clinical risk scores (CRS) that quantify bleeding risk can guide decision making in situations when indication or duration for anticoagulation is uncertain. We investigated whether integration of a genetic risk score (GRS) into an existing risk factor-based CRS could improve risk stratification for anticoagulation-related ICH.

**Methods:** We constructed 153 GRS from genome-wide association data of 1,545 ICH cases and 1,481 controls and validated them in 431 ICH cases and 431 matched controls from the population-based UK Biobank. The score that explained the largest variance in ICH risk was selected and tested for prediction of incident ICH in an independent cohort of 5,530 anticoagulant users. A CRS for major anticoagulation-related hemorrhage, based on 8/9 components of the HAS-BLED score, was compared with an enhanced score incorporating an additional point for high genetic risk for ICH (CRS+G).

**Results:** Among anticoagulated individuals, 94 ICH occurred over a mean follow-up of 11.9 years. Compared to the lowest GRS tertile, being in the highest tertile was associated with a two-fold increased risk for incident ICH (HR: 2.08 [95% CI 1.22, 3.56]). While the CRS predicted incident ICH with a HR of 1.24 per one point increase (95% CI [1.01, 1.53]), adding a point for high genetic ICH risk led to a stronger association (HR of 1.33 per one point increase, 95% CI [1.11, 1.59]) with improved risk stratification (C-index 0.58 vs. 0.53) and maintained calibration (integrated calibration index 0.001 for both). The new score (CRS+G) showed 20% improvement of high-risk classification among individuals with ICH and a net reclassification improvement of 0.10.

**Conclusions:** Among anticoagulant users, a prediction score incorporating genomic information is superior to a clinical risk score alone for ICH risk stratification and could serve in clinical decision making.

## Introduction

More than five million people in the United States use anticoagulants for prevention of ischemia or treatment of thrombosis.^1^ Anticoagulants effectively prevent thrombosis and thromboembolism, but at the same time also increase bleeding risk,^2^ of which intracerebral hemorrhage (ICH) is the most devastating outcome with a mortality of 30-50% and major disability among survivors.^3,4^ In situations when indication or duration for anticoagulation is uncertain,^5-9^ clinical risk scores (CRS) for major hemorrhage such as the widely used HAS-BLED score^10,11^ can help physicians weigh the individual bleeding risk against the expected benefits.^12,13^

Genetic risk scores (GRS) that leverage additive effects of common risk variants throughout the whole genome can provide risk stratification for complex diseases such as coronary artery disease and ischemic stroke with similar effect sizes as traditional cardiovascular risk factors.^14-17^ Individual genetic variation contributes substantially to ICH risk with a heritability of up to 44%,^18^ but this information has not yet been exploited for risk stratification and prevention. Recently, by aggregating genetic liability for risk factors underlying ICH, we were able to develop a GRS that was consistently associated with ICH in the general population.^19^ However, whether leveraging genetic variation that predisposes specifically to ICH risk could improve risk stratification and decision making among anticoagulant users remains unknown.

In the current study, we investigated whether genetic variation can be leveraged for ICH risk stratification among individuals on anticoagulants, a collective with known elevated risk for ICH. We hypothesized that with the currently available genome-wide association data, it is feasible to develop a GRS specific for ICH that associates with incident ICH among anticoagulant users. We furthermore hypothesized that a combination of clinical and genomic risk factors for prediction of ICH is superior to clinical risk factors alone.

## Methods

### UK Biobank

The UK Biobank (UKB) is an ongoing population-based prospective cohort study of 502,409 UK residents aged 40-69 years. Individuals were recruited from 2006-2010 in 22 assessment centers across the UK.^20^ Detailed baseline characteristics, a range of biochemical assays and genome-wide genotyping is available for most of the study participants. Genotyping was performed on two very similar platforms and imputation was performed on the Haplotype Reference Consortium for 488,377 participants.^21^

The UKB has institutional review board approval from the Northwest Multi-Center Research Ethics Committee (Manchester, UK). All participants provided written informed consent. We accessed the data following approval of an application by the UKB Ethics and Governance Council (Application No. 36993). We followed the recently published reporting standards for polygenic scores in risk prediction studies.^22^

### Derivation, validation, and test cohorts

The overall design of the study is depicted in **Figure 1**. The most recent ICH genome-wide association study (GWAS) meta-analysis from 1,545 ICH cases and 1,481 controls of self-reported European ancestry was used as derivation cohort to develop a GRS that predisposes to ICH risk.^23^ Summary statistics of effect estimates for variants imputed with 1000 Genomes reference panel were downloaded from the Cerebrovascular Disease Knowledge Portal (cd.hugeamp.org).

**Figure 1.**
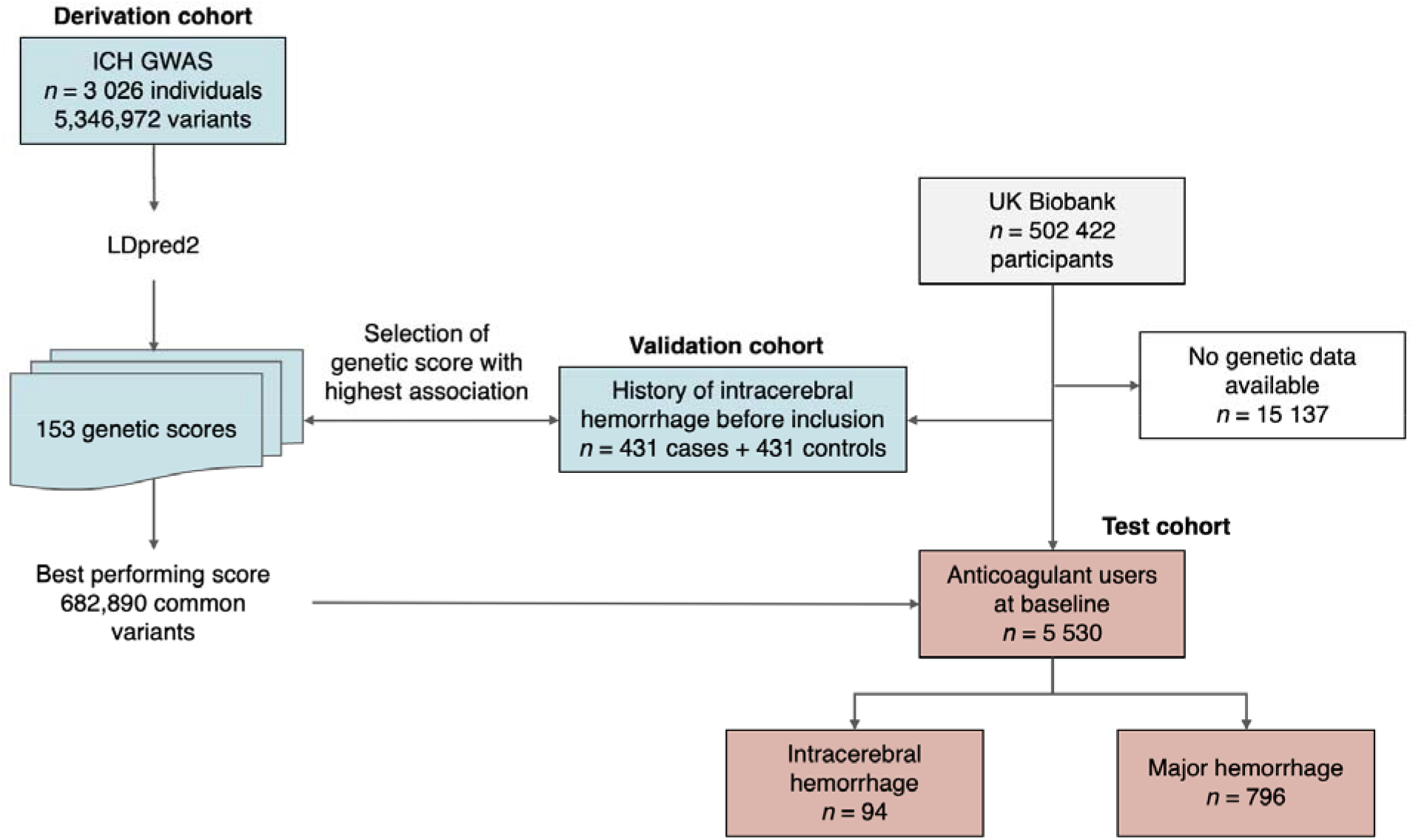
Study flow. Genetic risk scores for ICH were obtained from the most recent genome-wide association study (derivation cohort).^23^ Among participants with available genetic data, UK Biobank participants were grouped into the validation cohort that consisted of individuals with history of intracerebral hemorrhage (ICH) before inclusion, and the test cohort with individuals on anticoagulant intake at baseline. The score with the strongest association with ICH in the validation cohort was selected and tested on longitudinal ICH outcomes in the test cohort.

The validation cohort for the GRS consisted of UKB individuals of principal component (PC) derived European ancestry with history of ICH at baseline and the same number of randomly selected age- and sex-matched ICH-free controls.

The test cohort consisted of individuals without a history of ICH at baseline and anticoagulant use defined by self-report in the verbal interview at inclusion. Furthermore, individuals were included if they had a diagnosis of the International Classification of Diseases 10 (ICD-10) code Z92.1 (personal history of long-term (current) use of anticoagulants) or D68.3 (hemorrhagic disorder due to circulating anticoagulants) at baseline, or a prescription of an anticoagulant medication between baseline and six months thereafter in the primary care data (search terms supplied in **Supplementary Table S1**).

### Outcome ascertainment

The UKB provides medical history and ongoing longitudinal follow-up data by gathering inpatient hospital codes, primary care data, and death registry data. ICH and ischemic stroke events were assessed by the diagnostic algorithm for stroke in the UKB (https://biobank.ndph.ox.ac.uk/showcase/ukb/docs/alg_outcome_stroke.pdf) that captured inpatient hospital events by using outcome-specific ICD codes up to January 2022. Additional ICH events were gathered from the primary care data that captured events up to 2017 in 45% of participants (search terms supplied in **Supplementary Table S2**). Major hemorrhages were defined as bleeding events that required hospitalization including ICH, gathered from ICD codes of inpatient diagnoses (codes supplied in **Supplementary Table S3**). History of ICH was defined as ICH occurring before inclusion and incident events (ICH, ischemic stroke, major hemorrhage) were defined as events occurring after inclusion.

### Development and validation of the genetic score

LDpred2 was used with an external linkage disequilibrium (LD) reference panel as suggested by the authors^24^ with its three different modes LDpred2-inf, LDpred2-auto, and LDpred2-grid on a grid of hyperparameters (factor for *h2* of 0.3, 0.7, 1, and 1.4; sequence of *p* from 10^−5^ to 1; sparsity 0 and 1) to construct a total of 153 GRS (**Supplemental Table S4)**. Out of all derived scores, the score that explained the largest variance (highest Nagelkerke’s R^2^) for ICH in the validation cohort in logistic regression models adjusted for baseline age, sex, PCs 1-10, and genotyping array was selected. The final model for which effect estimates are reported was adjusted for age at baseline (controls) or ICH event (cases), sex, PC1-10, and genotyping array.

### Testing of the genetic score

The effect of the selected GRS on longitudinal outcomes in the test cohort was investigated in Cox proportional hazard models with ICH as outcome and the covariates age, sex, PCs 1-10, and genotyping array. Because of the low absolute number of ICH events, main analyses were performed in all individuals regardless of self-reported race, and sensitivity analyses were performed among cohorts stratified by self-reported race (white vs. non-white) and among unrelated individuals (KING kinship coefficient <0.0884). To investigate the specificity of the score, additional Cox models were constructed with the GRS as predictor, major hemorrhage and ischemic stroke as outcomes, and age, sex, PCs 1-10, and genotyping array as covariates. To evaluate whether the prediction power of the GRS varies by age or sex, interaction terms between the GRS and age and sex were modeled.

### Construction of a clinical and combined clinical and genetic risk score for major hemorrhage

We constructed a CRS mimicking HAS-BLED, a validated clinical score widely used for risk stratification for major hemorrhage among anticoagulant users.^10,11^ Verbal interview data, hospital inpatient diagnoses, and biochemical measures were leveraged to obtain the information required for the components of the CRS at baseline. Out of nine components, all but one (INR lability) were available. Similar to HAS-BLED,^10^ points were assigned for uncontrolled blood pressure, abnormal renal or liver function, prevalent stroke, bleeding history, age, and drug or alcohol intake. This CRS was then tested for its predictive capability in univariable Cox models on outcomes ICH and major hemorrhage.

To test whether the GRS contains specific genetic susceptibility for ICH beyond the information captured by the CRS, both the CRS and the PRS were used as independent predictor variables in a multivariable Cox model. In the next step, to enable a clinically useful tool in patients at high risk, we tested whether the continuous GRS could be simplified into a binary phenotype that indicates high risk for ICH. The optimal cut-off point was determined by maximizing the Youden’s index for classification of ICH events.^25^ A new score (CRS+G) was then constructed that included one additional point if an individual’s GRS exceeded the determined cut-off point. Univariable Cox proportional hazard models in the validation cohort were used to test the prediction capability of the CRS and the CRS+G to predict cause-specific subhazards for both incident ICH and major hemorrhage. Uno’s concordance index was used to evaluate the models’ discrimination and the integrated calibration index (ICI) to evaluate their calibration as recently suggested by the STRengthening Analytical Thinking for Observational Studies (STRATOS) initiative.^26^ To further evaluate the potential clinical utility of the score, the classification of individuals at high risk for bleeding was compared using the net reclassification improvement (NRI),^27^ using the cut-off of ≥ 3 points as previously suggested for HAS-BLED.^12,13,28^ All-time ICH or major bleeding events were used for calculation of NRI.

### Software used

For preparation of genotypic data, genetic score calculation, and relationship inference we used bcftools, plink, LDpred2, and KING.^29-32^ For data extraction, curation, preparation, and figure generation, we used RStudio 2022.07.0 with R version 4.2.1 on Mac OS X (aarch64-apple-darwin20) with the packages bigsnpr, coxphw, data.table, FSA, fmsb, gmodels, nricens, survival, survminer, survcomp, tidyverse, and writexl.^33^

### Data availability

The data that support the findings of this study are available from the UK Biobank upon submission of a research proposal.^20^ The summary statistics of the GWAS for ICH that was used for construction of the GRS are publicly available.^23^

## Results

### UK Biobank validation and test cohorts

The validation cohort consisted of 431 individuals with prevalent ICH and 431 age- and sex-matched controls with a mean age of 60 years and 41% females (**Table 1**). The test cohort consisted of 5,530 individuals on anticoagulants at baseline (2006-2010), mean age at baseline 59 years, 31% females). Most of the individuals in the test cohort (n=5,366, 97%) were on warfarin at baseline, other anticoagulants used were heparins, phenindione, and acenocoumarol (**Table 2**). 796 incident major hemorrhages including 94 incident ICH occurred over a mean follow-up of 11.9 years, and 82% of the individuals were censored after 10 years or later.

**Table 1.** Baseline characteristics of the validation cohort that consisted of individuals with history of ICH at baseline and randomly selected age- and sex-matched controls. Anticoagulant intake and other clinical risk factors at time of hemorrhage were unavailable from the ICH events that happened before inclusion in the UK Biobank.

| Validation cohort | ICH cases (n=431) | ICH-free controls (n=431) |
| --- | --- | --- |
| Age at inclusion, years, mean (SD) | 59.7 (7.1) | 59.7 (6.4) |
| Age at ICH, years, mean (SD) | 50.6 (11.1) | NA |
| Female, n (%) | 177 (41.2) | 178 (41.1) |
| European ancestry, n (%) | 431 (100) | 431 (100) |
| Genetic score, mean (SD) | -0.68 (0.37) | -0.62 (0.39) |
| Genetic score highest tertile*, n (%) | 168 (39.0) | 131 (30.3) |

\* tertiles of the genetic scores were derived among all UK Biobank participants with genetic data ICH, intracerebral hemorrhage

**Table 2.** Baseline characteristics of the test cohort that consisted of 5,530 individuals on anticoagulants at baseline.

| **Baseline characteristics** | **ICH-free participants**<br>**(n=5,436)** | **Participants with incident ICH (n=94)** |
| --- | --- | --- |
| Age, years, mean (SD) | 61.9 (6.4) | 63.6 (5.1) |
| Female, n (%) | 1,695 (31.2) | 27 (28.7) |
| Warfarin intake, n (%) | 5,275 (97.0) | 91 (96.8) |
| Atrial fibrillation, n (%) | 2,827 (52.0) | 62 (66.0) |
| European ethnicity, n (%) | 4,972 (91.5) | 86 (91.5) |
| Follow-up duration, years, mean (SD) | 11.9 (3.3) | 6.9 (3.9) |
| **Clinical risk score components** |  |  |
| Uncontrolled hypertension, n (%) | 591 (10.9) | 14 (14.9) |
| Abnormal renal function n (%) | 54 (1.0) | 1 (1.1) |
| Abnormal liver function n (%) | 5 (0.1) | 0 (0) |
| History of stroke, n (%) | 634 (11.7) | 16 (17.0) |
| Prior major bleeding or bleeding predisposition, n (%) | 524 (9.6) | 7 (7.4) |
| Age > 65 years, n (%) | 1,889 (34.7) | 43 (45.7) |
| Alcohol or drug usage (≥ 8 drinks/week), n (%) | 2,178 (40.1) | 31 (33.0) |
| Medication usage predisposing to bleeding, n (%) | 795 (14.6) | 20 (21.3) |
| **Score values** |  |  |
| Clinical risk score (CRS), median (IQR) | 1 (1-2) | 1 (1-2) |
| Genetic score, mean (SD) | -0.65 (0.39) | -0.58 (0.37) |
| Highest genetic risk*, n (%) | 1,934 (35.6) | 46 (48.9) |
| Clinical and genetic risk score (CRS+G), median (IQR) | 1 (1-2) | 1 (1-3) |
| **Outcomes** |  |  |
| Incident major hemorrhage, n (%) | 714 (13.1) | 94 (100) |
| Incident ischemic stroke, n (%) | 301 (5.5) | 24 (25.5) |

\* cut-off for highest genetic risk was determined by maximizing Youden's index

### Derivation of the best GRS for ICH

The best-performing GRS in the derivation cohort was the one with the highest R^2^ (0.02) and the lowest p value (0.012) and was obtained from LDpred2-grid with parameters *h2*=0.7288, *p*=0.1, and *sparse*=*FALSE* incorporating information from 682,890 common genetic variants. Among all UKB participants, the genetic score had a mean of -0.66 and a standard deviation (SD) of 0.39 (**Figure 2A**). One SD of the score increased the odds for ICH by an OR of 1.30 in the validation cohort (95% CI [1.11, 1.52]) without evidence of non-linearity (**Figure 2B)**. The weights of the GRS are supplied in **Supplemental Table S5**.

**Figure 2.**
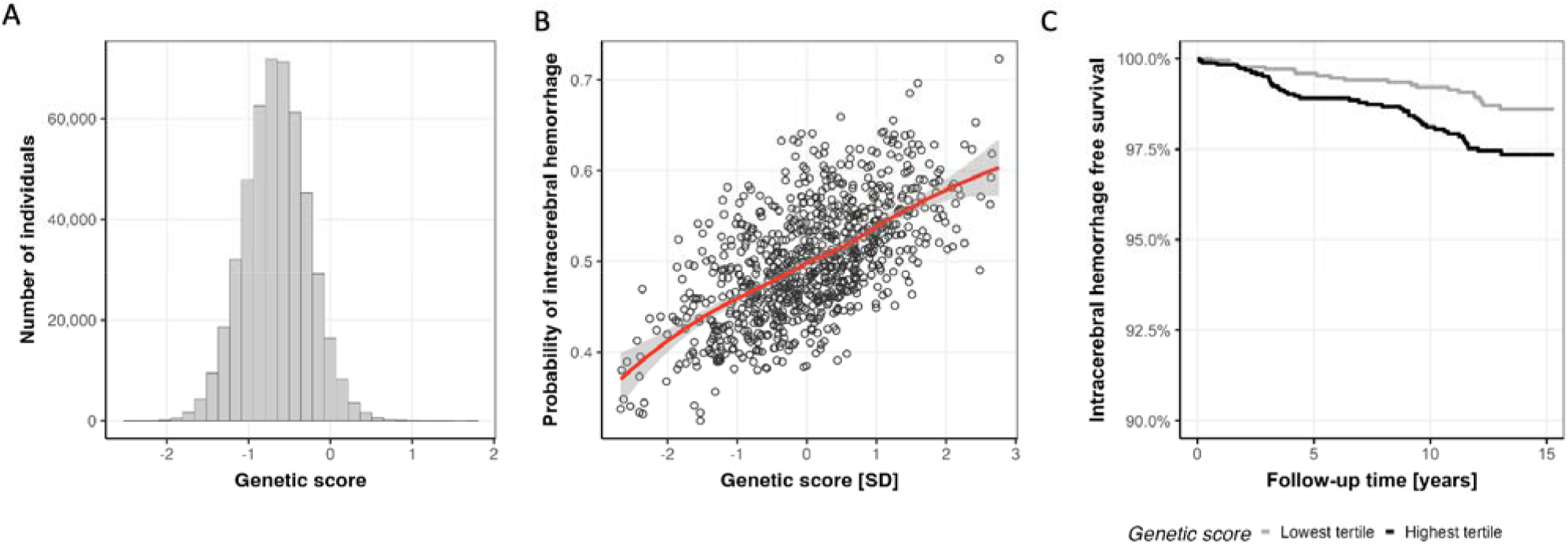
Genetic risk score (GRS) for intracerebral hemorrhage (ICH). **A**: Distribution of the obtained GRS for ICH. **B**: The GRS for ICH was significantly associated with prevalent ICH in the derivation cohort without evidence of nonlinearity. **C**: The polygenic risk for ICH through common genetic variation among anticoagulant users is substantial: after adjusting for age, sex, and hypertension, the hazard ratio for ICH was 2.08 (95% CI [1.22, 3.56]) for individuals in the highest tertile of the GRS compared to the lowest tertile.

### Associations of the GRS with incident ICH

The selected GRS was significantly associated with incident ICH (HR of 1.24 per one SD, 95% CI [1.01, 1.54]) in the test cohort. Compared to the lowest tertile of the score, individuals in the highest tertile had a HR of 2.08 for ICH (95% CI [1.22, 3.56]; **Figure 2B**). Sensitivity analyses confirmed the association in the test cohort restricted to individuals of European ancestry (n=4,818, HR 1.75, 95% CI [1.01, 3.04]) and non-European ancestry (n=472, HR 28.8, 95% CI [1.56, 533.31]). The association was also confirmed among unrelated individuals of the test cohort (n=5,139, HR 1.89, 95% CI [1.10, 3.26]). There was no association of the GRS with incident major hemorrhage (p=0.88) and incident ischemic stroke (p=0.82), and no interaction between the genetic score and age or sex (both p>0.3).

### Added value of the genetic score on top of a clinical risk score for major hemorrhage

The CRS for major hemorrhage mimicking HAS-BLED had a median of one point and 519 (9.4%) individuals had more than two points (**Figure 3A**). Similar to HAS-BLED,^10,11^ one CRS point increase was associated with a higher risk for incident major hemorrhage (HR=1.18, 95% CI [1.09, 1.26]) and risk for incident ICH (HR=1.24, 95% CI [1.01, 1.53], **Figure 3C**). The GRS was associated with incident ICH independent of the CRS (HR 1.24 per one SD increase, 95% CI [1.01, 1.54]), indicating genetic susceptibility for ICH beyond captured clinical risk factors.

**Figure 3.**
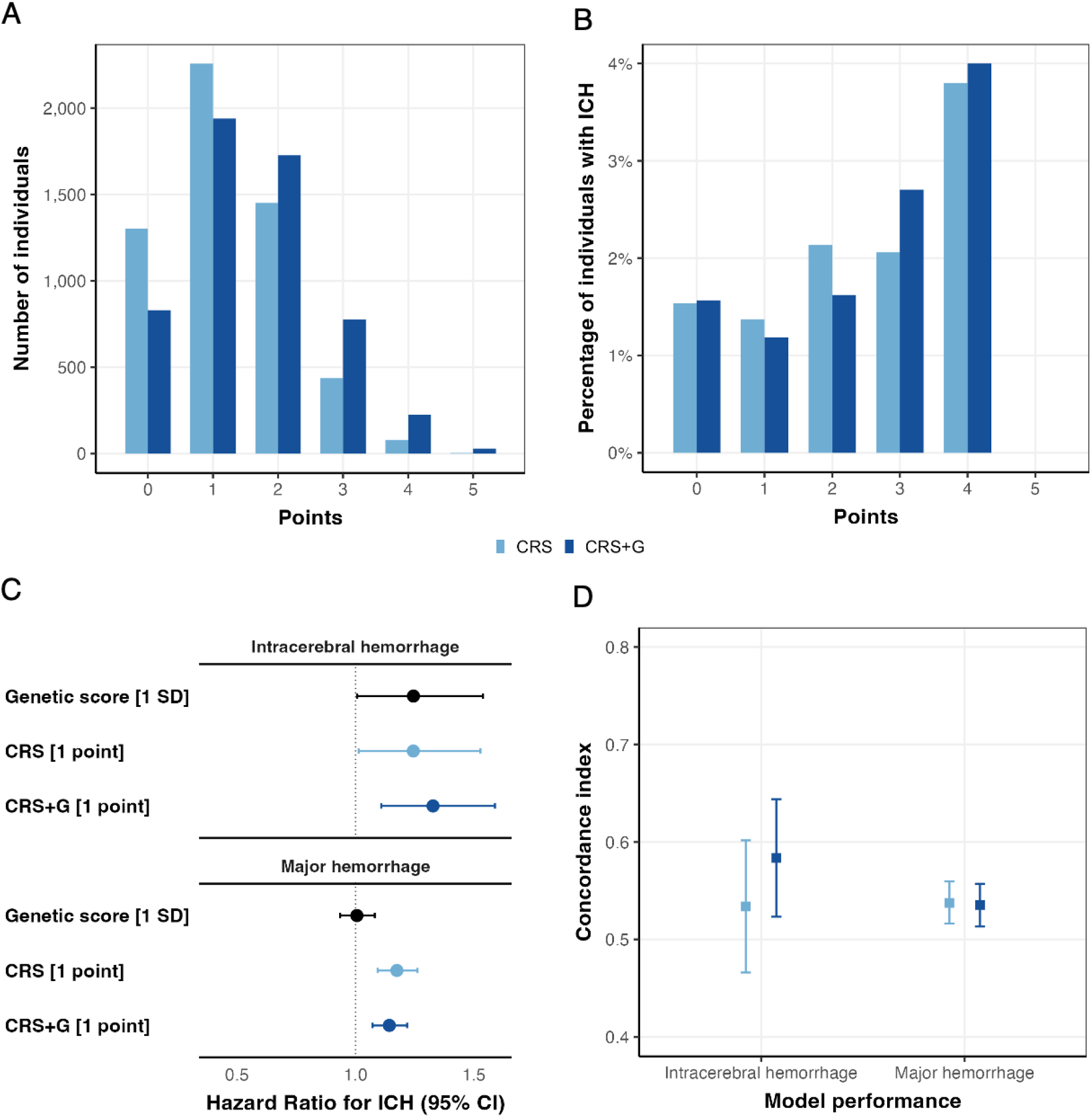
The combination of clinical and genetic risk factors has the strongest prediction effect on intracerebral hemorrhage among 5,530 anticoagulant users over mean 11.9 years of follow-up (test cohort). **A**: Distribution of the clinical risk score (CRS) compared to the combined clinical and genetic risk score (CRS+G) in individuals with anticoagulants at baseline. **B**: Percentage of individuals suffering ICH per score point of the CRS and the CRS+G. **C**: Both the genetic risk score and the clinical risk score were only borderline significantly associated with incident ICH. Combination of the genetic and clinical score into the CRS+G yielded a robust association with incident ICH, while maintaining a similar association with any major hemorrhage including ICH. **D**: The CRS+G had a better discrimination for 10-year risk of incident ICH than the CRS, while maintaining similar discrimination for any major hemorrhage as the CRS.

To reduce the information content of the continuous GRS for potential clinical utility, we investigated whether incorporation of high genetic susceptibility for ICH would improve the discriminative power of the CRS while retaining its bedside utility. Thus, one point was added to the CRS for individuals with a GRS of more than -0.057 (n=1,980, 35.8%), the cut-off value with the highest Youden’s index for classifying individuals at high genetic risk. The resulting CRS+G significantly increased the risk for incident ICH with a HR of 1.33 (95% CI [1.11, 1.59]) while maintaining its association with incident major hemorrhage (HR 1.14, 95% CI [1.07, 1.22], **Figure 3C**). Accordingly, the all-time risk of suffering an ICH increased more monotonically for increasing points of the CRS+G compared to the CRS alone (**Figure 3B**).

Discrimination and calibration were compared for the prediction of events at ten years, a timepoint when the majority (82%) of the individuals at risk had follow-up data. The discrimination of the CRS+G for incident ICH was higher (c-index 0.58, 95% CI [0.52,0.64]) compared to the CRS (c-index 0.53, 95% CI [0.47, 0.60]), while the discrimination of major hemorrhage remained similar (c-index 0.54, 95% CI [0.52,0.56] vs. 0.54 95% CI [0.51,0.56], respectively, **Figure 3D**). The calibration was very similar for both scores (ICIs for CRS 0.001 and 0.11 and for CRS+G 0.001 vs. 0.11 for ICH and major hemorrhage, respectively).

Finally, we compared the CRS and CRS+G in terms of their classification of individuals at high risk for bleeding (≥ 3 points). For incident ICH, the NRI was 0.10 (95% CI 0.04, 0.15), the sum of the 19% improvement in classification as high-risk patients of those who experienced an ICH (NRI_E_) at the cost of 9% more of ICH-free individuals classified as high-risk (NRI_NE_). For major hemorrhage the NRI was 0.02 (95% CI 0.00, 0.04), the sum of 11% improvement in classification of high-risk patients (NRI_E_) at the cost of 9% more patients classified as high-risk who never experienced a bleeding event (NRI_NE_).

## Discussion

Using data from the largest currently available ICH GWAS, we leveraged LDpred2 to construct a well-performing genetic risk score for ICH among prevalent cases and controls in the UK Biobank.^23,31^ We validated the predictive effect on incident ICH in a separate cohort of 5,530 anticoagulant users with a follow-up of 12 years. We further showed that integration of genetic risk for ICH into a CRS based on the HAS-BLED score improves its discrimination for ICH while maintaining its association with major hemorrhage. When comparing the classification of high-risk individuals, the CRS+G showed a net reclassification improvement for both ICH and major hemorrhage, with a 20% improvement for classifying high-risk individuals for ICH.

Our study demonstrates the utility of a GRS in a clinically relevant population. Although many GRS have been developed and validated in the general population to predict diseases, they still face challenges including portability and interpretability before application in routine clinical care.^34^ GRS could be especially useful in high-risk populations to identify individuals with additional high genetic risk for intervention trials.^35^ Anticoagulant users are at increased ICH risk, and the additional knowledge of individual genetic ICH risk could influence clinical decision making in situations with limited evidence or equivocality of clinical parameters. For potential cardioembolic sources of ischemic stroke such as left ventricular aneurysm,^8^ ventricular thrombus,^36^ or low ejection fraction,^37^ anticoagulation indications are controversial, but also for well-established indications such as deep venous thrombosis, pulmonary embolism, and cerebral sinus thrombosis, duration of anticoagulation is uncertain and provoked versus unprovoked status can be ambiguous.^5-9^ Randomized controlled trials validating bleeding risk scores are missing, but the clinical utility is demonstrated by the mentioning of HAS-BLED in American and European guidelines.^12,13^ When using a fixed threshold for classification of individuals at high risk for hemorrhage as suggested for the clinical setting,^12,13^ our CRS+G prioritizes ICH risk, at the cost of putting 9% of individuals who might never experience an ICH in a higher risk category. Incident ICH events are infrequent compared to all major hemorrhages (in our test cohort 94 vs. 796 events over 12 years) and have thus been underrepresented in the development of HAS-BLED.^10,11^ But ICH is the most feared adverse event of anticoagulants with enormous morbidity and mortality^38^, affecting patients more than other hemorrhages that have a smaller impact on quality-adjusted life years^39^ and hence it might be reasonable to prioritize superior high-risk classification for ICH over that for any major hemorrhage. Thus, in situations where the question arises whether anticoagulation should be withdrawn, our CRS+G could be leveraged to weigh bleeding risk against thrombosis or thromboembolism risk.

The combined clinical-genetic risk score was superior to a solely clinical risk score for prediction of ICH. Traditional risk scores evaluating all major hemorrhages are not optimized for ICH, but because the heritability of ICH by common genetic variation is high (up to 44% overall; 73% for lobar and 34% for non-lobar)^18^, genetic instruments can provide robust risk estimates. Larger GWAS could provide a more robust and generalizable GRS, but due to the rarity of ICH, GWAS efforts have been slower than for ischemic stroke.^23,40^ Despite these challenges, our results demonstrate that in a score of eight different clinical risk factors for major hemorrhage, one additional point for the highest genetic ICH risk substantially improves the prediction of incident ICH. Thus, in situations with clinical equipoise, we submit that individual genetic ICH risk could be accounted for when stratifying an individual both for ICH and major hemorrhage risk.

Our study has limitations. First, the constructed GRS was derived from an ICH GWAS of individuals of European ancestry, because this is currently the largest available genome-wide association data.^23^ Even though the GRS performed well among European and non-European ancestry individuals, a more diverse training set could improve performance. Thus, future research should focus in particular on underserved populations that have higher burden and earlier onset of ICH.^41^ Larger studies to better characterize the genetic architecture of ICH in diverse populations are underway.^42^ Second, the vast majority (97%) of patients were on warfarin because that was the dominant oral anticoagulant at the time of recruitment into the UK Biobank, thus the findings cannot be translated to direct oral anticoagulants, some of which have lower reported hemorrhage risk. However, because only ∼20% of the ICH GWAS cases used for GRS derivation were on anticoagulants^23^ and because our genetic score was selected in the validation cohort among prevalent ICH cases agnostic to anticoagulation status, it was not specifically designed for performance in anticoagulant-associated ICH and may therefore applicable to other high-risk situations. Moreover, in many clinical settings, warfarin remains the first choice, such as for rheumatic heart disease associated atrial fibrillation,^43^ prosthetic heart valve and antiphospholipid syndrome, and in patients with severe liver or kidney disease. Third, because of apparent power problems in the existing data, we were not able to show meaningful decision curve analysis or net reclassification data in comparison with risk scores for ischemic stroke. Future releases of outcome data will allow finer analyses to help in balancing ischemic versus hemorrhagic stroke risk. Finally, the absolute risk for ICH, even for individuals on anticoagulation was low, calling into question the clinical relevance of this relative risk increase. However, given the lack of treatment options for ICH, its excessive morbidity and mortality, and the substantial role of genetic liability for its occurrence, genetic risk stratification could still be clinically useful in the emerging era of more cost-effective genotyping.

In conclusion, a prediction score incorporating genomic information is superior to a clinical risk score alone for ICH risk stratification and could aid in clinical decision making among anticoagulant users. While this approach has a high potential for clinical utility, future studies with larger sample sizes of more diverse subjects are warranted to confirm our findings. Our results could inform secondary analyses of existing trial data and future trial design in order to demonstrate and deliver individualized treatment in the era of precision medicine.

## Data Availability

The data that support the findings of this study are available from the UK Biobank upon submission of a research proposal. The summary statistics of the GWAS for ICH that was used for construction of the GRS are publicly available.

